# An MRI-Based Staging System for Osteochondritis Dissecans Demonstrates Substantial Interrater Reliability and Tracks Progressive Ossification During Healing

**DOI:** 10.1101/2025.05.08.25326569

**Authors:** Brent Burg, Rohan Raikar, Michael Newcome, Abdul Wahed Kajabi, Eisa Hedayati, Saumith Bachigari, Karsten Knutsen, Shelly Marette, Joe Luchsinger, Takashi Takahashi, Kevin Klee, Jesse Reiter, Marc Tompkins, Lin Zhang, Jutta Ellermann

## Abstract

**Purpose:** To evaluate the interrater reliability and clinical applicability of a novel MRI-based radiological staging system for osteochondritis dissecans (OCD) that incorporates a short echo time GRE sequence to assess progressive ossification and healing status.

**Methods and Materials:** This retrospective HIPAA-compliant study was approved by the institutional IRB. MRI exams from April 2017 to December 2023 were reviewed for patients undergoing diagnostic OCD evaluation. Inclusion required the first MRI with a short echo time GRE (Gradient-Recalled Echo) and TSE (Turbo Spin Echo) sequences. Fifty-two MRIs (mean patient age: 13.4 ± 3.8 years; 28 male, 24 female knees) were randomly selected to ensure balanced stage distribution. Five musculoskeletal radiologists and fellows independently applied the proposed staging system based on progressive ossification, bridging, and lesion stability. Interrater reliability was measured using Fleiss’ Kappa. Healing outcomes were stratified as: (i) early surgery, (ii) successful non-operative therapy, or (iii) delayed surgery after failed non-operative management. Mean healing times were compared across groups using ANOVA.

**Results:** Substantial interrater reliability (Fleiss’ Kappa = 0.71, 95% CI: 0.65–0.77; p < 0.01) indicates strong agreement across five readers. Among 43 cases with clinical data, 19% (n=8) underwent immediate surgery, while 81% (n=35) received initial non-operative care; **29%** (n=10) later required surgery. Healing times differed significantly (p = 0.002, ANOVA): 0.75 ± 0.38 years for early surgery, 0.86 ± 0.62 years for successful non-operative treatment, and 2.4 ± 1.5 years for failed non-operative management with delayed surgery. Findings support the reproducibility of the staging system and its potential to identify lesions at risk of failed non-operative healing.

**Conclusion:** This novel MRI-based radiological staging system demonstrates substantial interrater reliability and enables tracking of progressive ossification, improving assessment of OCD healing. It’s integration of short echo time GRE sequences supports broader application in musculoskeletal imaging, including monitoring of fracture healing.

**Key Results:** *High Interrater Reliability:* The novel MRI-based Osteochondritis Dissecans (OCD) staging system demonstrated substantial interrater reliability (Fleiss’ Kappa = 0.71), supporting its reproducibility for clinical and research use.

*Healing Time Differentiation:* Healing timelines differed significantly across treatment groups—patients with failed non-operative therapy required nearly three times longer to heal than those who underwent early surgery or successfully completed conservative treatment.

*MRI-based Ossification Tracking with broader Applicability:* The staging system effectively visualized progressive ossification using short echo time GRE sequences (TE < 2.6 ms), enabling assessment of healing stages not captured by conventional MRI sequences. Beyond OCD, this framework may be applicable to other conditions involving endochondral ossification, such as fracture healing.

**Summary Statement:** A novel MRI-based radiological staging system for osteochondritis dissecans demonstrates substantial interrater reliability and enables assessment of healing through improved visualization of progressive ossification using short echo time GRE.

## INTRODUCTION

Osteochondritis Dissecans (OCD) is a developmental disease primarily affecting the knees in children and adolescents with an incidence of 6-10 per 100,000 and a significantly higher rate in boys^1^. OCD is characterized by focal failure of endochondral ossification, increasing the risk of developing loose bodies, and early-onset osteoarthritis^2–5^. The historical differentiation between juvenile and adult OCD has been reconsidered, as emerging consensus recognizes adult OCD as a continuum of the same developmental disease^6^. Common symptoms include pain, swelling, and locking, significantly impacting physical activity and well-being in young, active individuals^2, 7^.

The etiology of OCD is multifactorial, involving epiphyseal cartilage necrosis^8^, biomechanical overload^9^, and genetic predisposition^2, 7, 10^. These factors disrupt endochondral ossification, leading to focal zones of arrested bone formation at the osteochondral junction^3, 4^.

Importantly, successful OCD healing recapitulates endochondral ossification—beginning with epiphyseal cartilage necrosis and a delayed ossification front (Stage I), followed by peripheral rim mineralization (Stage II), internal ossification and partial bridging (Stage III), and ultimately complete osseous integration (Stage IV). Notably, this biologic cascade mirrors fracture healing and skeletal repair, both of which proceed through these sequential stages^11^. Ellermann et. al. developed an MRI-based staging system incorporating a short echo time GRE sequence (TE < 2.6 ms), enabling improved contrast for detecting progressive ossification^4^. This facilitates reliable assessment of osseous healing and overcomes limitations of conventional Turbo Spin Echo (TSE) sequences (TE ≥ 20 ms), which cannot reliably distinguish ossified from non-ossified tissues^12^.

Current MRI classification systems focus on lesion stability^13–15,16^, effectively guiding treatment for only 5 – 15 % of lesions that are clearly unstable. However, the majority of lesions are classified as stable, yet 25–67 %^17–20^ of these ultimately require surgery. These staging systems cannot track ossification because they solely rely on TSE sequences, resulting in poor interrater reliability. This contributes to limited diagnostic accuracy^21^ and variability in patient care^2, 18, 19^.

We hypothesize that an MRI-based radiological staging system aligned with the natural ossification trajectory of OCD will demonstrate high interrater reliability and provide clinically relevant imaging findings for assessing healing – offering a framework applicable to imaging-guided assessment of musculoskeletal repair.

## MATERIALS and METHODS

### Population

The University of Minnesota IRB approved this retrospective, HIPAA-compliant study and waived the need for informed consent. This study included MRIs obtained at the University of Minnesota, Fairview from April 28, 2017, to December 28, 2023, for the diagnostic workup of OCD lesions. Inclusion criteria required the imaging study to be the patient’s first MRI for OCD including a Gradient recalled echo (GRE) T2* sequence.

Initially, 198 consecutive MRI studies were available, but 94 did not meet the inclusion criteria (Figure 1)_. Among excluded cases, alternative diagnoses included control knees (n=40), post-surgical changes (n=21), ossification variants (n=22), and chondromalacia (n=3). Exclusion ensured cohort specificity. The remaining 104 studies met the inclusion criteria for pre-surgical OCD (64 males, 40 females; mean age 13.8 ± 4.2 years). Each case was initially assigned a stage (Figure 1) by a fellowship trained MSK radiologist, 15 years of clinical and research expertise in OCD (JE). From each stage, approximately half of the cases were randomly selected for inclusion (See Figure 1, Post Stratification and Randomization) using Microsoft Excel to assign case numbers. For Stage II lesions with initially 29 cases, where the midpoint equaled 14.5, the number was rounded down; for Stage IV with initially 5 cases, the number was rounded up to ensure adequate representation. This resulted in 52 cases—half of the full cohort—chosen to balance statistical rigor with practical feasibility for a multi-reader study. This sample size exceeded the minimum of 22 cases determined by power analysis to achieve 95% statistical power with five raters. Importantly, this subset ensured a balanced distribution across all stages. After a secondary image quality review, six cases were exchanged with the next in line randomly assigned study within the same stage ensuring that all included cases were complete and met image quality standards.

**Figure 1.**
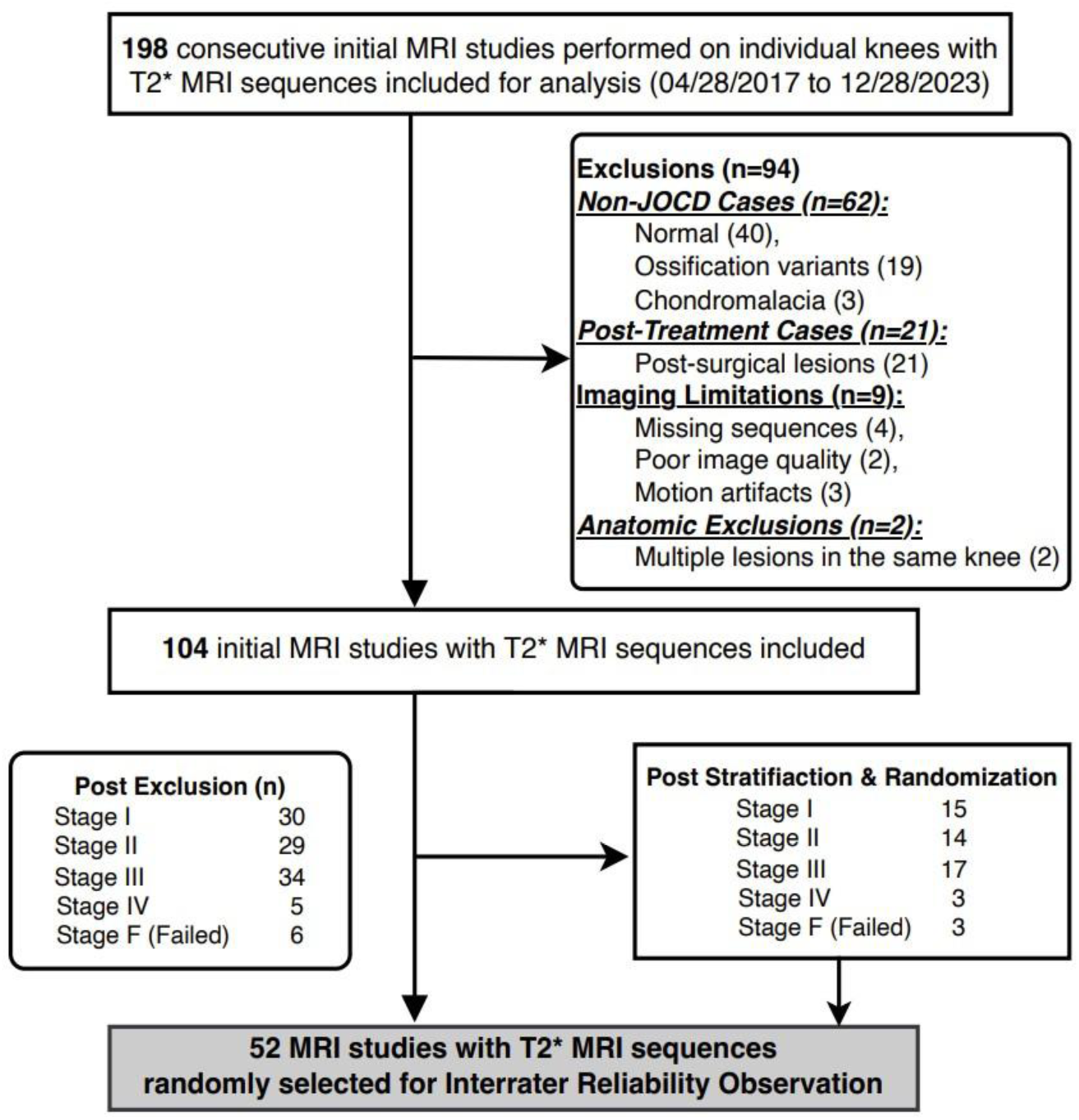
Exclusion Criteria: Flowchart displays the selection criteria, timeline and exclusion criteria. Studies were assigned a randomly generated number within stratified categories of participant’s sex, knee laterality and initial stage.

### Clinical Data Analysis

Clinical outcome was defined as lesion healing. Healing (test positivity) was defined as either (1) complete resolution of clinical symptoms and/or (2) evidence of complete osseous healing on follow-up MRI. These categories were pre-specified based on clinical guidelines and prior imaging studies. Healing could occur with: (i) early surgery, (ii) successful non-operative therapy, or (iii) delayed surgery after failed non-operative treatment. Partial healing on MRI or ongoing symptoms were classified as non-healing. This composite clinical and imaging reference standard was selected over arthroscopy, as the latter was not uniformly available and therefore not used as a reference standard. Assessors of the reference standard, the clinical outcome data had full access to patient medical records, including clinical presentation, treatment history, and imaging. However, they were blinded to the test results of the OCD staging to prevent interpretation bias.

The initiation of therapy was determined using the date of first documented treatment in the electronic medical record (EMR). Healing was defined by either MRI confirmation of Stage IV (complete osseous integration) and/or clinical resolution of symptoms. For each group, the mean time and standard deviation from therapy initiation to documented healing were calculated.

### Imaging Protocol(s)

All 52 MRI studies were collected at our institution at 3T (Prisma Fit, Siemens Healthineers, Erlangen, Germany) using a birdcage transmit with a 15-channel receive array coil, or 7T (Terra, Siemens Healthineers, Erlangen, Germany) using a birdcage transmit with a 28-channel receive array coil. The applicability of these findings is not limited by field strength, as T2* imaging provides high SNR across different field strength. While our studies were acquired at 3T and 7T, the underlying imaging principles and clinical utility remain broadly applicable to all field strength, including 1.5T. The MRI protocol included routine clinical T1-, T2- and PD-weighted multi-slice TSE sequences acquired at both 3T and 7T systems (Table 1). In addition, short echo time 2D GRE or 3D GRE sequences were obtained using TE of 2.6 at 3T and 1.4 ms at 7T, respectively. Images from the shortest attainable echo time of the GRE sequence were displayed with inverted contrast, resembling computed tomography (CT)-like bone contrast.

**Table 1.**
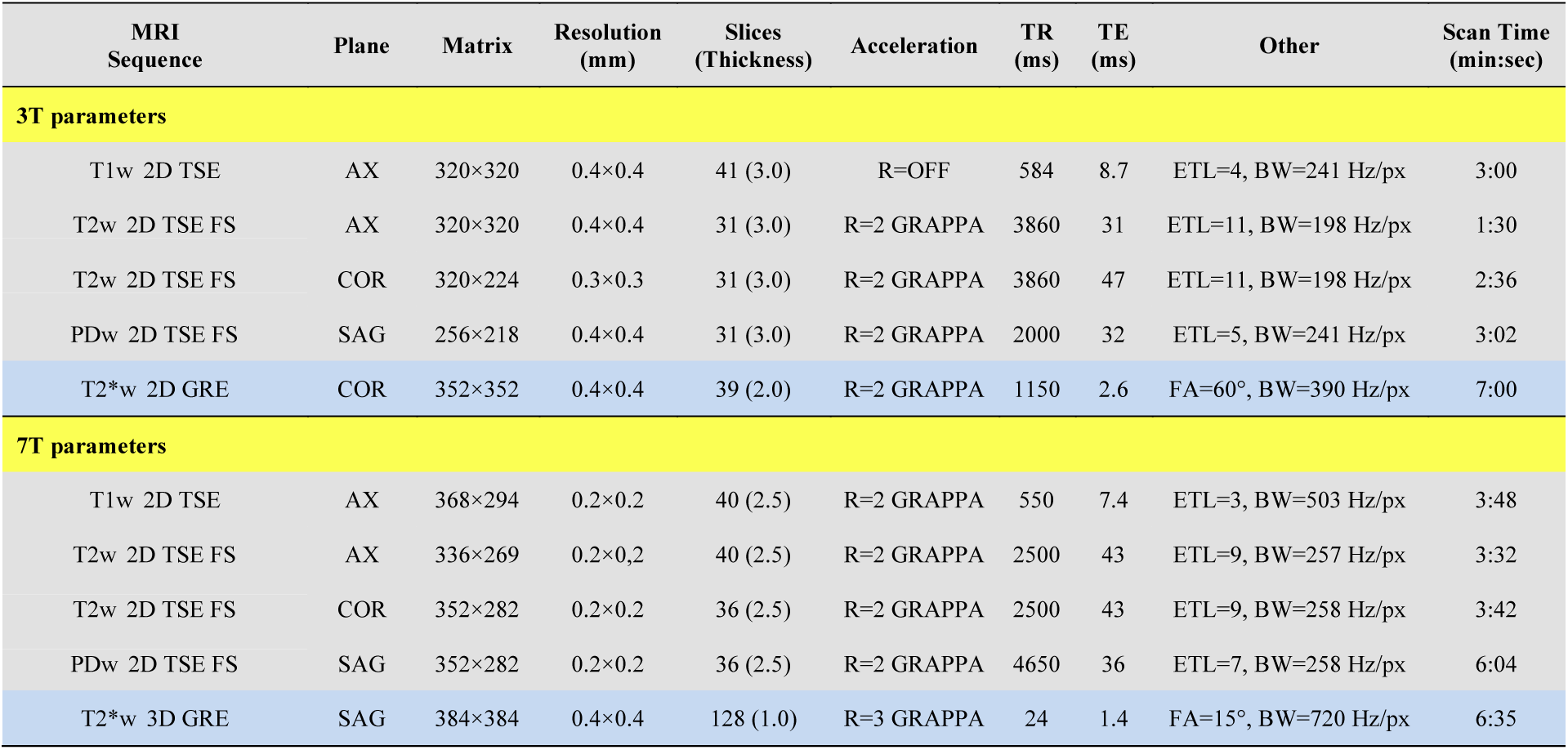
MRI sequence parameters. TR=repetition time; TE= echo time; AX=axial; COR=coronal; SAG=sagittal; TSE=turbo spin echo; GRE=gradient-recalled echo. Grey: Clinical TSE sequences; Blue: CT-like bone contrast.

| MRI Sequence | Plane | Matrix | Resolution (mm) | Slices (Thickness) | Acceleration | TR (ms) | TE (ms) | Other | Scan Time (min:sec) |
| --- | --- | --- | --- | --- | --- | --- | --- | --- | --- |
| **3T parameters** |  |  |  |  |  |  |  |  |  |
| T1w 2D TSE | AX | 320×320 | 0.4×0.4 | 41 (3.0) | R=OFF | 584 | 8.7 | ETL=4, BW=241 Hz/px | 3:00 |
| T2w 2D TSE FS | AX | 320×320 | 0.4×0.4 | 31 (3.0) | R=2 GRAPPA | 3860 | 31 | ETL=11, BW=198 Hz/px | 1:30 |
| T2w 2D TSE FS | COR | 320×224 | 0.3×0.3 | 31 (3.0) | R=2 GRAPPA | 3860 | 47 | ETL=11, BW=198 Hz/px | 2:36 |
| PDw 2D TSE FS | SAG | 256×218 | 0.4×0.4 | 31 (3.0) | R=2 GRAPPA | 2000 | 32 | ETL=5, BW=241 Hz/px | 3:02 |
| T2*w 2D GRE | COR | 352×352 | 0.4×0.4 | 39 (2.0) | R=2 GRAPPA | 1150 | 2.6 | FA=60°, BW=390 Hz/px | 7:00 |
| **7T parameters** |  |  |  |  |  |  |  |  |  |
| T1w 2D TSE | AX | 368×294 | 0.2×0.2 | 40 (2.5) | R=2 GRAPPA | 550 | 7.4 | ETL=3, BW=503 Hz/px | 3:48 |
| T2w 2D TSE FS | AX | 336×269 | 0.2×0.2 | 40 (2.5) | R=2 GRAPPA | 2500 | 43 | ETL=9, BW=257 Hz/px | 3:32 |
| T2w 2D TSE FS | COR | 352×282 | 0.2×0.2 | 36 (2.5) | R=2 GRAPPA | 2500 | 43 | ETL=9, BW=258 Hz/px | 3:42 |
| PDw 2D TSE FS | SAG | 352×282 | 0.2×0.2 | 36 (2.5) | R=2 GRAPPA | 4650 | 36 | ETL=7, BW=258 Hz/px | 6:04 |
| T2*w 3D GRE | SAG | 384×384 | 0.4×0.4 | 128 (1.0) | R=3 GRAPPA | 24 | 1.4 | FA=15°, BW=720 Hz/px | 6:35 |

### Staging System Validation

Five reviewers, three board certified musculoskeletal radiology attendings (SM, JL, TT), and two musculoskeletal radiology fellows independently staged the studies. Experience with OCD lesions varied widely among the readers from the fellows with one year of experience in musculoskeletal radiology and occasional exposure, two attendings with 15 and 8 years of experience having only occasional exposure, and one attending with 10 years of experience and extensive exposure. All imaging studies were de-identified, independently and sequentially reviewed by readers who were blinded to clinical information and reference standard outcomes. Reviewers received general instructions on 10 pilot cases. Representative imaging features for each stage of healing and failed healing, are illustrated in Figures 1–5. Readers opened studies to the predetermined most representative slice and staged lesions based on the representative slice plus or minus one slice on either side. The most representative slice was determined to have the most advanced characteristic within a given lesion. For example, in a lesion that had a component that was entirely cartilaginous (stage I) but another region that showed rim mineralization (stage II), the slice with rim mineralization was chosen. This highlights the progressive nature of the stages and disease process, except in stage IV where the lesion must be completely ossified and bridged to parent bone. The MRI-based OCD staging system and imaging protocol are described in sufficient detail to allow full replication in clinical and research settings and ensure reproducibility in accordance with STARD guidelines.

**Figure 2.**
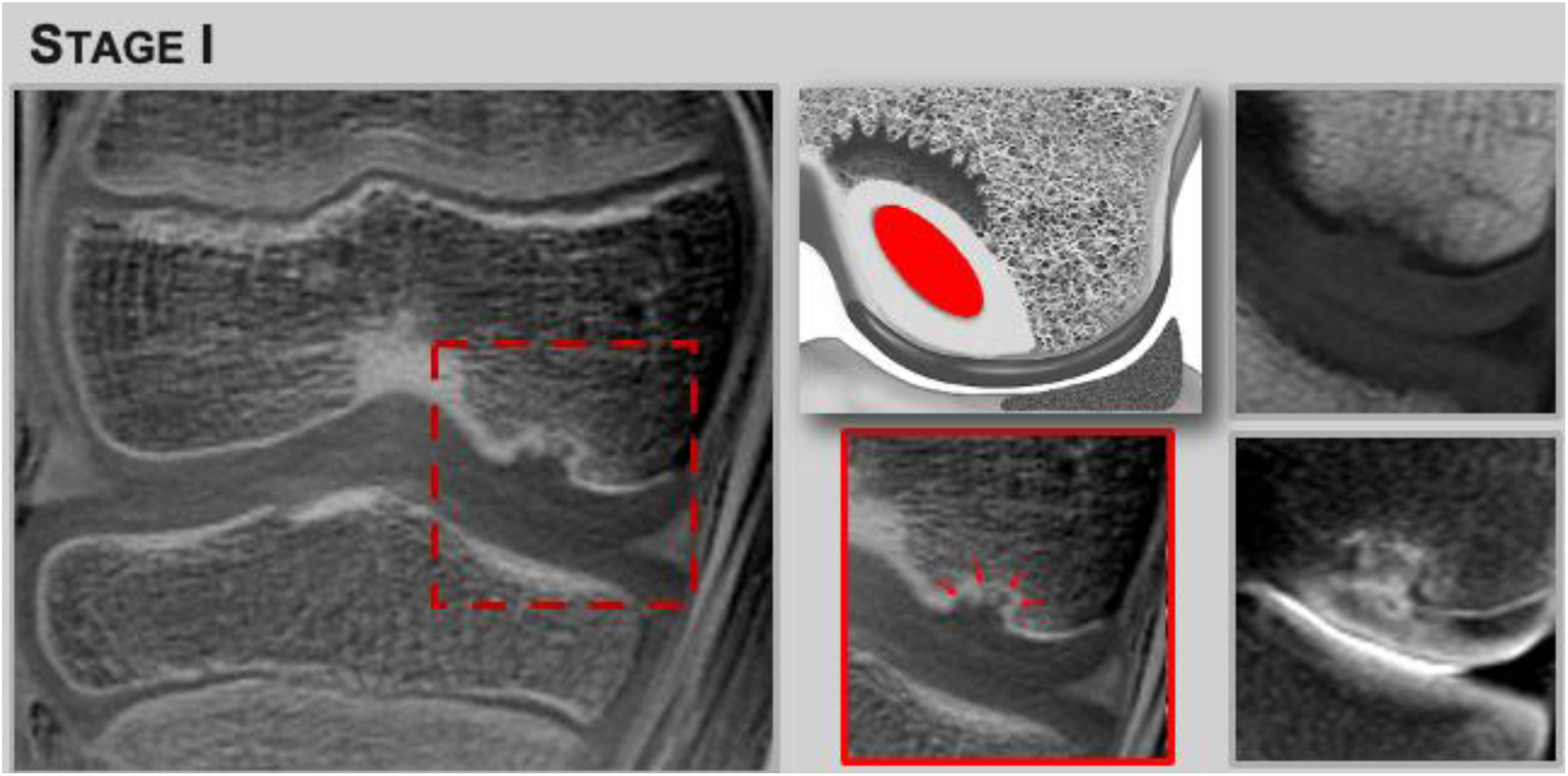
Stage I: Epiphyseal Cartilage Origin with Delayed Ossification. ***Left:*** T2*-GRE (short echo time, inverted contrast) shows a delayed ossification front and absence of internal lesion ossification. ***Middle:*** Schematic (top) illustrates epiphyseal cartilage origin of the lesion (red); T2* insert (bottom) highlights delayed ossification with characteristic “shark tooth” appearance (red arrows). ***Right:*** PD non–fat-saturated (top) and T2 FS TSE (bottom) provide limited visualization of the ossification front and tissue characterization of the lesion.

**Figure 3.**
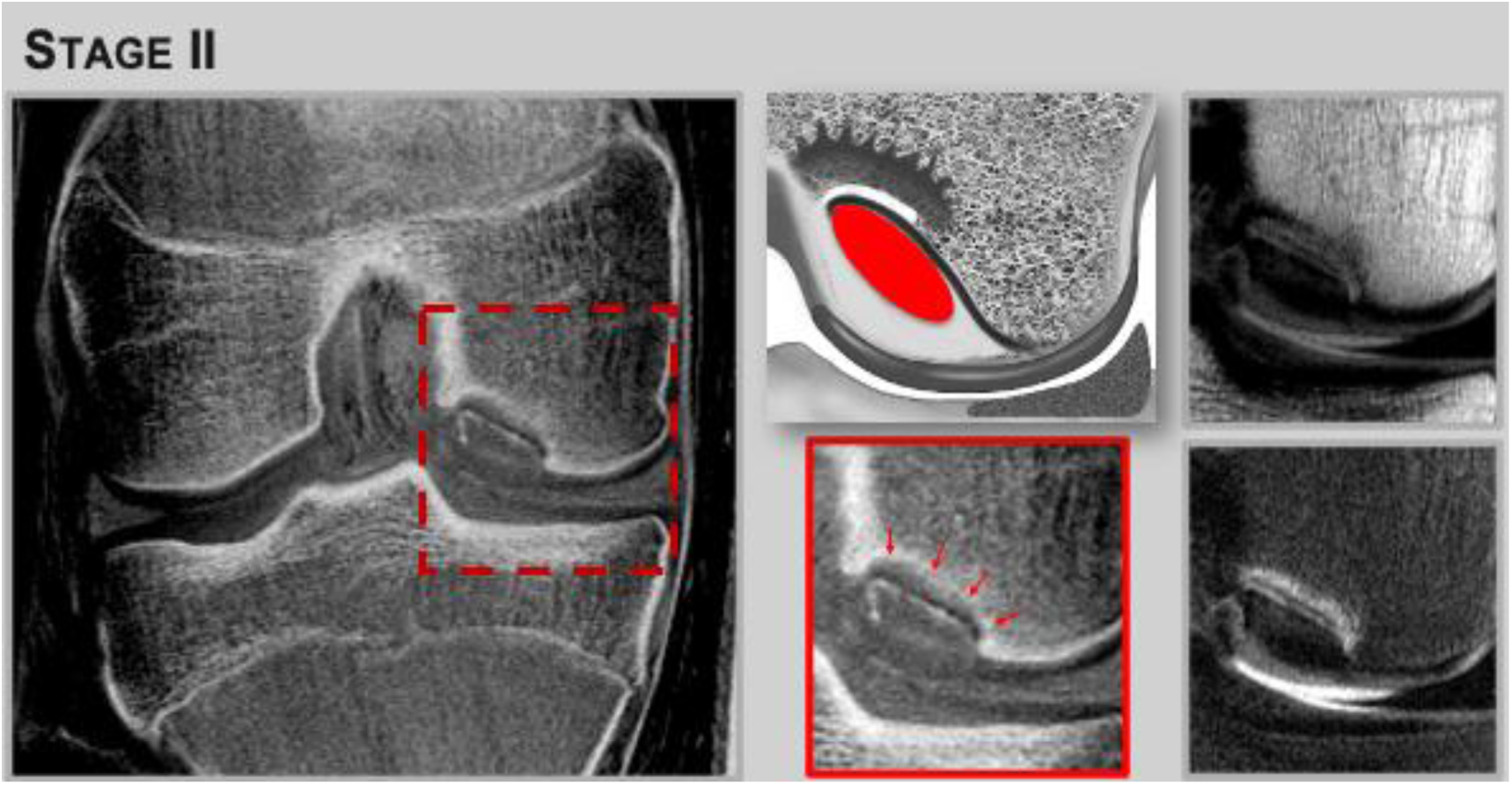
Stage II: Peripheral Rim Mineralization – First Step in Ossification. ***Left:*** T2*-GRE (short echo time, inverted contrast) shows peripheral rim mineralization of the lesion. ***Middle:*** Schematic (top) illustrates rim mineralization (black line) initially surrounding the lesion at the parent bone - lesion interface; T2* insert (bottom) highlights the delayed ossification front (red arrows) and the lesion peripheral rim mineralization. ***Right***: PD non–fat-saturated (top) and T2 FS TSE (bottom) provide limited information on tissue composition; the lesion remains fibrocartilaginous with a rim of mineralization. Due to long echo times, both cortical bone and fibrocartilage appear hypointense with lack of contrast, as all spins are fully relaxed.

**Figure 4.**
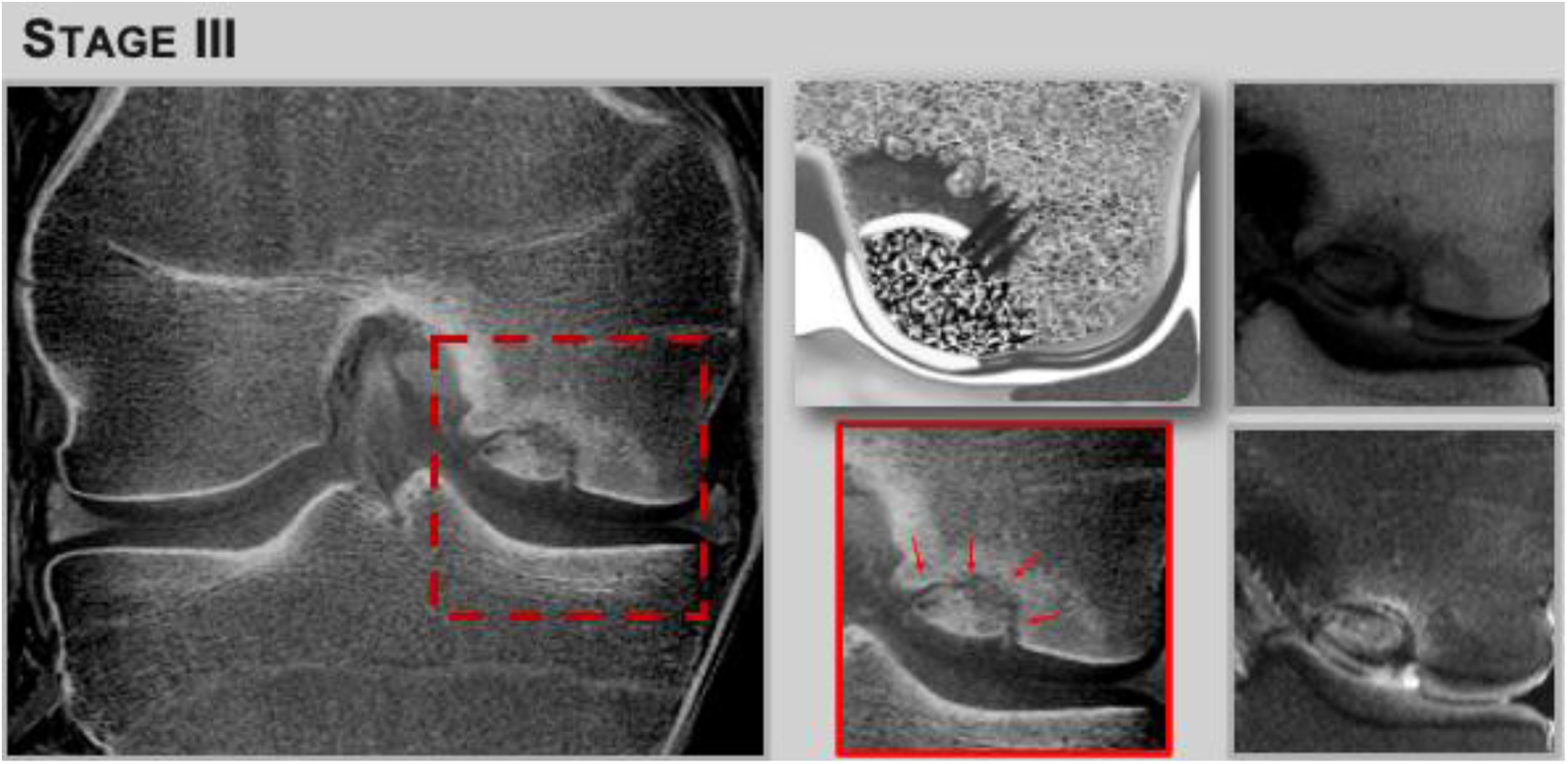
Stage III: Lesion Ossification – Transitional Phase of Ossification and Bridging. ***Left:*** T2*-GRE (short echo time, inverted contrast) shows progressive internal ossification within the lesion. ***Middle:*** Schematic (top) illustrates the ossified lesion and early bridging; T2* insert (bottom) highlights the parent bone -lesion interface (red arrows) with partial osseous bridging. ***Right:*** PD non– fat-saturated (top) shows absence of normal fatty marrow within the lesion and adjacent bone; T2 FS TSE (bottom) demonstrates intralesional edema. Both sequences lack clear identification of tissue composition, specifically osseous tissue due to long echo times.

**Figure 5.**
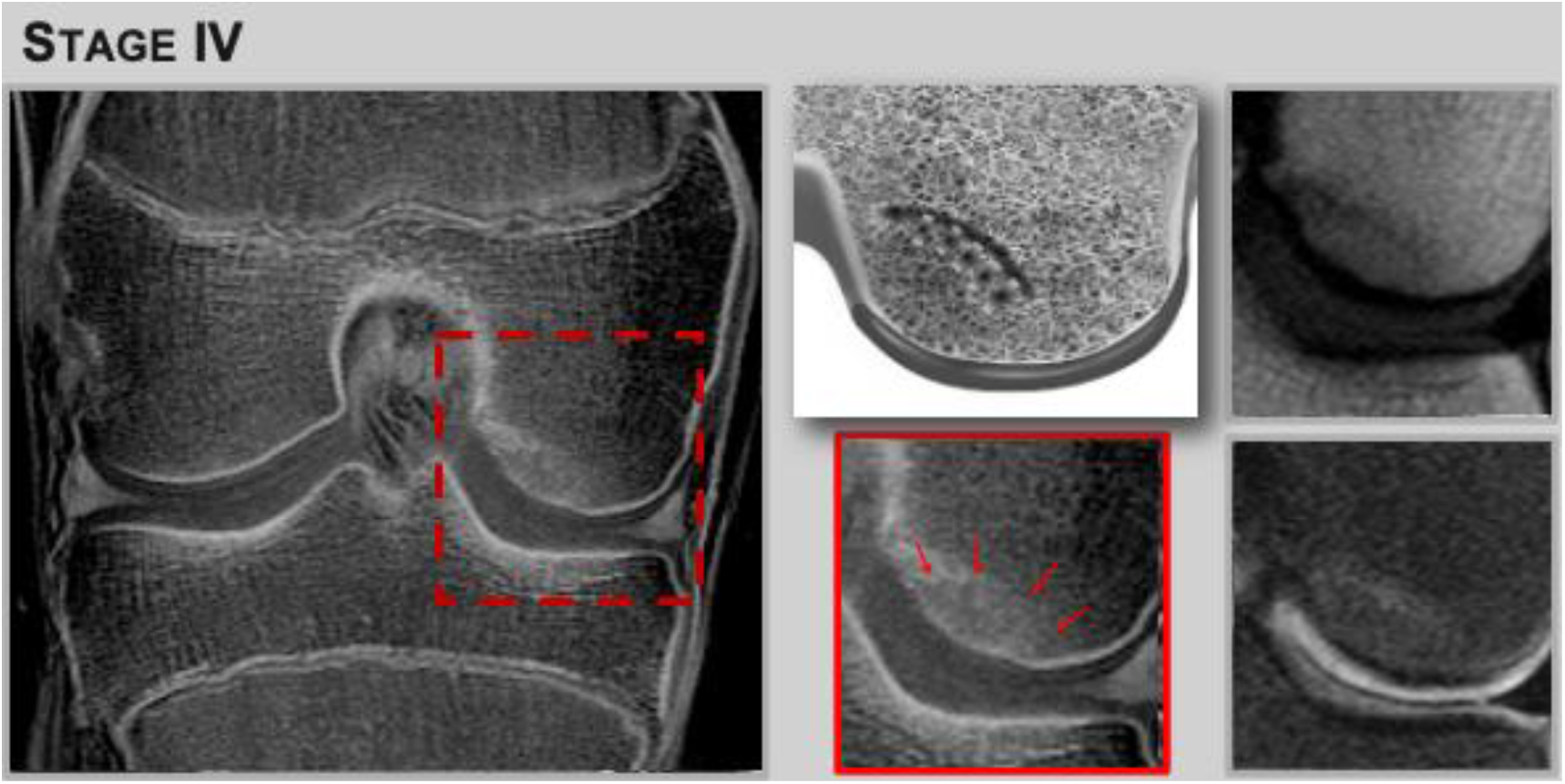
Stage IV: Complete Ossification and Bridging – Final Stage of Healing. ***Left:*** T2*-GRE (short echo time, inverted contrast) shows full lesion ossification and bridging with the parent bone. ***Middle:*** Schematic (top) illustrates complete integration; T2* insert (bottom) highlights the fully ossified interface (red arrows). ***Right:*** PD non–fat-saturated (top) depicts a subtle hypointense line consistent with the “scar sign“ at the healed the lesion – bone interface; T2 FS TSE (bottom) shows a faint “halo” of edema around the lesion.

### Statistical Analysis^22^

The power calculation using the ‘kappaSize’ R package indicates that with 5 raters, 52 cases have over 95% power to detect a tested kappa value of 0.8 compared to a null value of 0.5, at significance level of 0.05.

Our primary outcome measure was the interrater reliability of the staging calculated via Fleiss’ Kappa on rBiostatistics.com (Fleiss’ Kappa | rBiostatistics.com. Accessed July 16, 2024. https://rbiostatistics.com/node/67) with assistance from a statistician (LZ). The level of agreement was interpreted on a scale of 0.00-1.00, with 0.00-0.20 indicating poor agreement, 0.40-0.60 moderate agreement, 0.60-0.80 substantial agreement and 0.80-1.00 almost perfect agreement. Statistical significance was defined as a p-value of less than 0.05.

Mean healing times and standard deviations were calculated for immediate surgery, successful non-operative therapy and failed non-operative therapy groups. Statistical comparisons were performed using one-way ANOVA, followed by Tukey HSD post-hoc analysis to assess significant differences in healing duration between groups. Kruskal-Wallis testing, a non-parametric alternative, was also applied to account for variance heterogeneity across groups.

## RESULTS

Fifty-two knees were analyzed (mean age 13.4 ± 3.8 years). Of these, 28 were male knees and 24 female knees, with three participants contributing both knees. Substantial agreement was observed for the primary outcome of interrater reliability (Kappa = 0.71, 95% CI: 0.65–0.77; p < 0.01), supporting consistent and reproducible lesion staging.

Based on blinded rater staging via this system, stage II lesions (n = 16) were the most frequently identified, followed by stage III (n = 15), stage I (n = 13), Stage F (n = 4), and stage IV (n = 3). One lesion did not reach a consensus.

Of the 52 knees, three studies lacked sufficient clinical data for timeline analysis, and six participants were still undergoing treatment. As a result, 43 knees (21 female and 22 male knees, average age of 13.7 years ± 4.1) had complete clinical timelines available for analysis. Of these, 19% (n=8) required immediate surgery based on lesion instability on MRI, significant clinical symptoms, or patient preference for early intervention, with MRI instability being the only objective criterion. The remaining 81% (n=35) were managed initially with non-operative therapy. Among the latter group 71% (n=25) healed successfully with non-operative therapy, while 29% (n=10) ultimately required delayed surgical intervention. Mean healing times varied significantly between treatment groups (Figure 7). Patients who underwent early surgery healed in an average of 0.75 years (273 + 138 days). Those who healed with non-operative therapy alone had a comparable mean healing time of 0.86 years (313 + 227 days). Finally, patients whose stable lesions failed non-operative treatment and required delayed surgery averaged 2.4 years (882 + 560 days) before healing.

**Figure 6.**
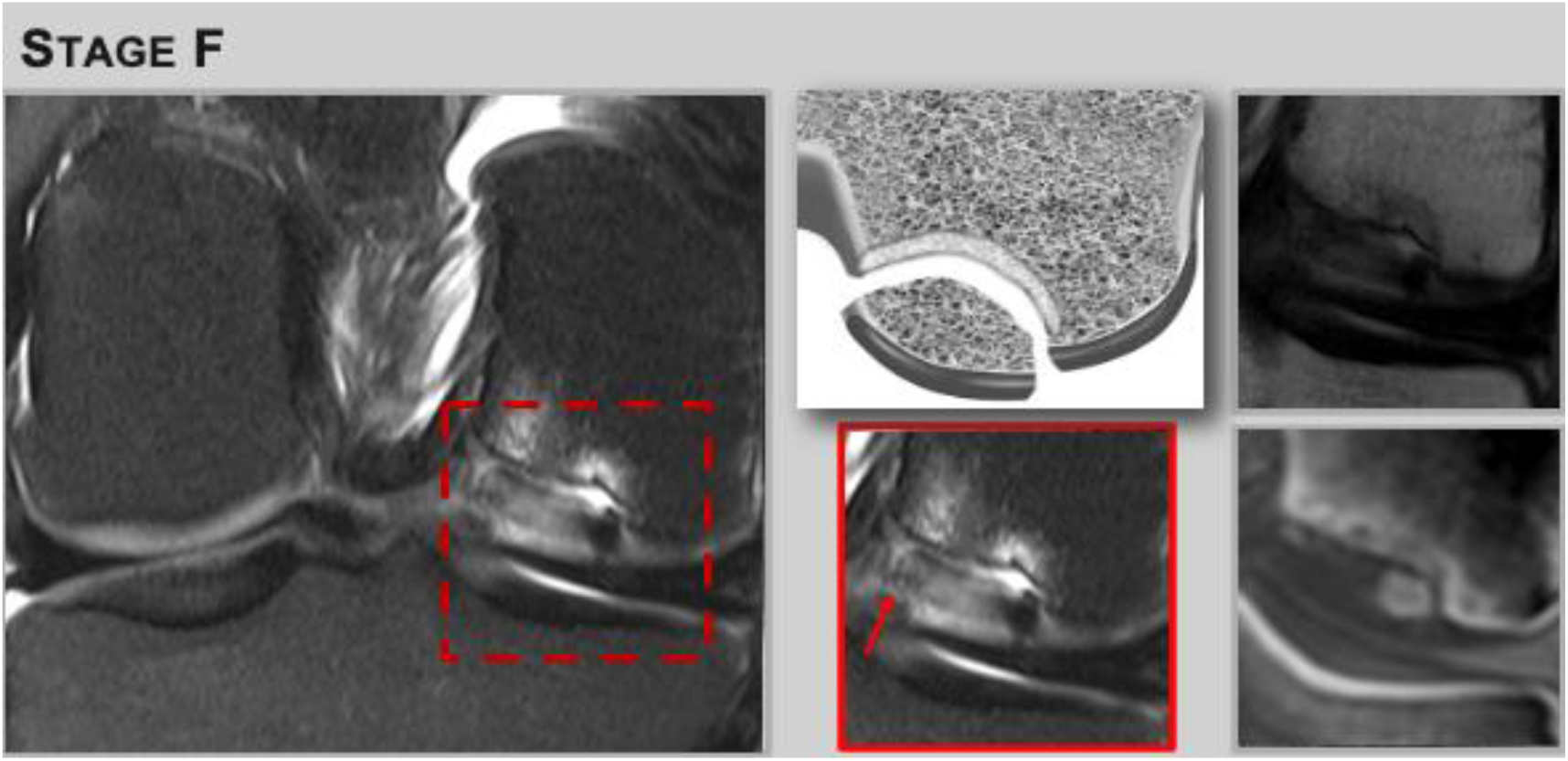
Stage F: Failed Healing – Instability and Lack of Progressive Ossification and Bridging. ***Left:*** T2 FS TSE reveals fluid signal at the parent bone–lesion interface and an articular cartilage fissure, indicating lesion instability. This sequence provides the primary diagnostic criteria for failed healing and should be reviewed first in image interpretation. ***Middle*:** Schematic (top) depicts failed healing which can lead to loose body formation; T2 FS TSE insert (bottom) highlights interface fluid and cartilage fissure (red arrow). ***Right:*** PD non–fat-saturated TSE (top), no information on lesion tissue composition; T2*-GRE (bottom) demonstrates partial internal lesion ossification without bridging, supporting the diagnosis of failed healing. T2*-GRE is used after stability has been assessed to determine the lesion’s tissue composition.

**Figure 7.**
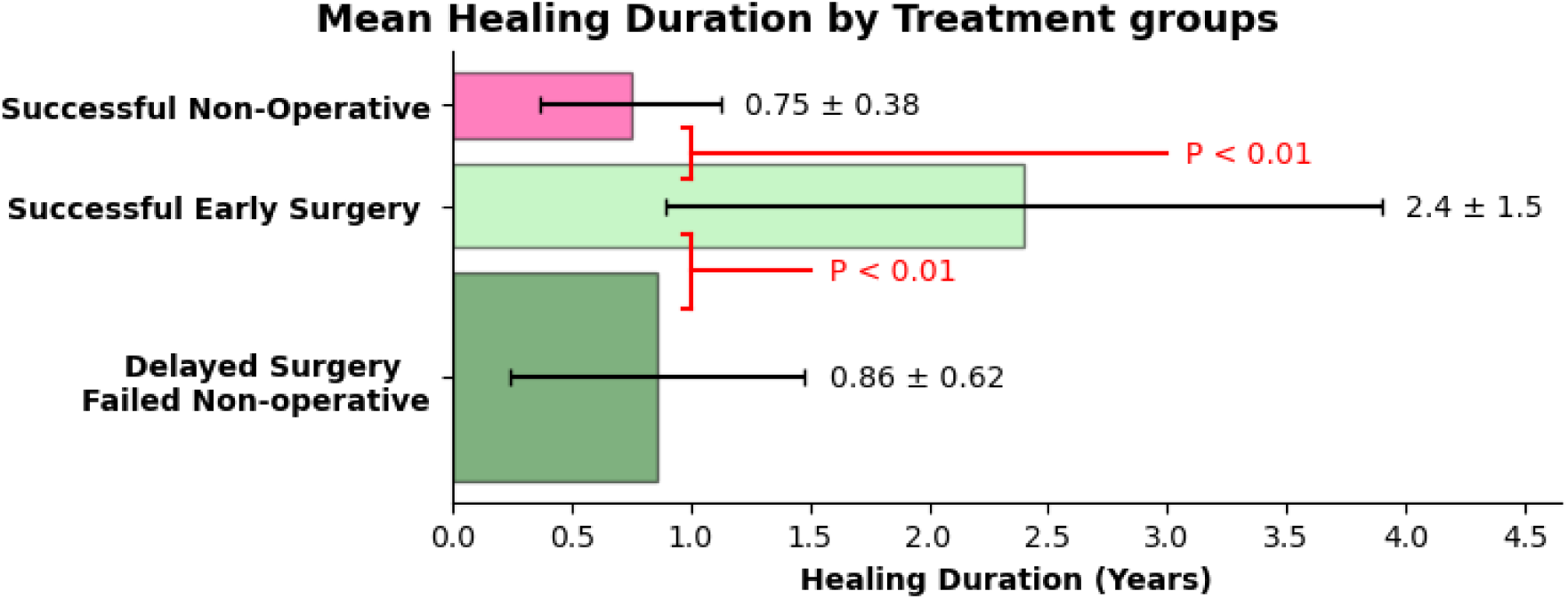
Bar graph showing mean healing durations in years with standard deviation for three patient groups: successful early surgical intervention; n=8 (mean 0.75 ± 0.38 years), the majority of patients n=25 successfully healed with non-operative treatment; (mean 0.86 ± 0.62 years), and delayed surgery after failed non-operative treatment; n=10 (mean 2.4 ± 1.5 years). Time to healing was significantly longer in patients whose lesions failed non-operative therapy (p < 0.01) highlighting the clinical importance of early identification of non-healing lesions.

A One-Way ANOVA demonstrated statistically significant differences in healing times across the treatment groups (p = 0.002). Tukey HSD post-hoc analysis confirmed significant differences between the failed non-operative group and both the successful non-operative group (p = 0.002) and the immediate surgery group (p = 0.009), but not between the immediate surgery and successful non-operative therapy (p = 0.948), which was also supported by Kruskal-Wallis testing (p = 0.019).

## DISCUSSION

This study demonstrates substantial interrater reliability (Fleiss’ Kappa = 0.71, 95% CI: 0.65–77) for a novel MRI-based staging system of OCD, which aligns with the biological sequence of ossification. By capturing ossification front delay, peripheral mineralization, lesion ossification plus bridging, and full integration, this system reflects the natural healing trajectory. Unlike traditional MRI classification systems that focus primarily on stability, this approach incorporates the continuum of healing stages, addressing a major limitation in existing OCD staging systems.

### Limitations of Stability-Based Staging Systems and Clinical Implications

Current MRI classification systems are essential for determining initial treatment—surgical versus non-operative—based on lesion stability on MRI and clinical symptoms^23^. However, stability alone lacks prognostic value. While 19% of our cohort required immediate surgery, one third (29%) of stable lesions eventually failed non-operative treatment. These findings align with literature reports that 25-67% of stable lesions ultimately require surgery^17–20^. Importantly, delayed surgical intervention in these patients significantly prolonged healing times compared to those who healed successfully with non-operative treatment or underwent early surgery.

These outcomes underscore the need for a reproducible system that assesses healing potential, not just lesion stability. Our system offers this by classifying lesions according to the degree of ossification and bridging—providing clinicians with a more precise evaluation of tissue composition, such as the distinction between fibrocartilage and mineralized bone, rather than generalized assessments of lesion stability. This staging becomes especially powerful when applied longitudinally, as most clinical protocols already include a follow-up MRI, usually after 3 months. Without an objective framework, follow-up assessments often remain vague, describing lesions as “unchanged” or “stable” without informing clinical action. The proposed system enables objective tracking of ossification progression, potentially improving the timing of surgical intervention.

### Contextualizing with Prior Classification Systems

Earlier classification systems—radiographic, MRI-based, and arthroscopic—each present limitation. Radiographs are insensitive for early diagnosis and stability evaluation^6, 13, 24^. MRI-based systems such as those by de Smet ^25,34^, Hefti^26^ and Kijowski^14^ et al., emphasized fluid signal at the interface, subchondral cysts, and cartilage integrity to define stability, but did not account for ossification status. Interrater reliability among these systems remains modest Dipaola^24,26,15^, reporting Krippendorf alpha of 0.5, 0.51 and 0.46, respectively. The arthroscopic grading system developed by the ROCK group^27, 28^ improved intraoperative assessment but is invasive and not applicable for follow up MRI-evaluations.

In contrast, our system leverages biologically grounded stages of healing and MRI signal contrast to standardize assessments. This framework allows clinicians to move beyond static assessments of stability and adopt a dynamic understanding of healing progression.

### Imaging Technique and Rationale

A critical innovation of this system is the incorporation of short echo time GRE sequences. Conventional TSE MRI lacks the contrast to differentiate fibrous and fibrocartilaginous from ossified tissues due to long echo times (Figure 8). GRE sequences (TE ∼1.5–3 ms) captures signals from all musculoskeletal tissues except tightly bound spins from bone^29, 30^ producing so-called “black bone” or inverted CT-like contrast that highlights ossification^31^. GRE sequences are widely available in clinical MRI protocols, and the T2*-weighted GRE used here is compatible with standard imaging platforms. Other advanced techniques like ultra-short echo time (UTE) MRI offer enhanced bone contrast^32^ but are not widely implemented. Therefore, our system can be rapidly adopted without requiring specialized software or hardware.

**Figure 8.**
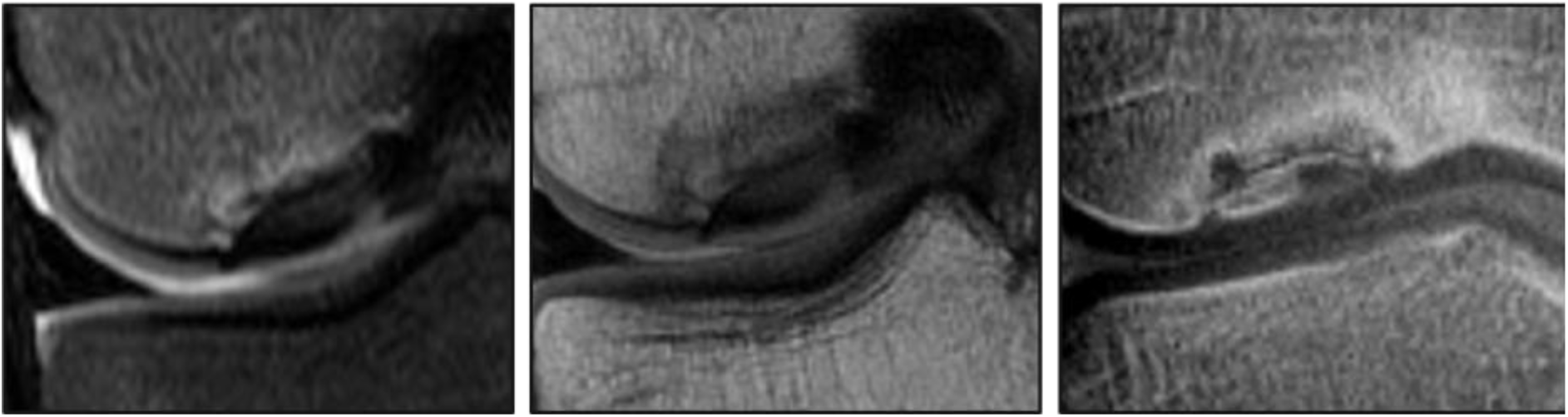
Coronal images illustrating the impact of echo time (TE) on lesion tissue differentiation. (Left) T2-weighted fat-saturated TSE (TE 47 ms) and (Middle) PD-weighted non-fat-saturated TSE (TE 32 ms): both fail to differentiate lesion tissue or the lesion-interface, as components such as mineralization, ossification, and fibrocartilage appear uniformly dark. (Right) CT-like GRE (TE 2.6 ms) with inverted contrast where bone appears bright. This enables clear identification of lesion ossification, allowing improved assessment of healing stage. The lesion is a stage III lesion, which reveals a largely ossified lesion without significant bridging at the interface between the parent bone and the lesion. There is parent bone sclerosis.

### Application to Clinical Decision-Making

The clinical implications of healing-based staging are considerable. In our study, healing times were comparable between patients who underwent early surgery and those who completed non-operative therapy (∼0.75 and 0.86 years, respectively). In contrast, stable lesions that failed non-operative treatment healed in 2.4 years—nearly three times longer. This highlights the importance of early identification of non-healing lesions and underscores the need for imaging frameworks that inform treatment timing. Current clinical guidelines by the *American Academy of Orthopaedic Surgeons* remain inconclusive, with significant variability in OCD management and healing timelines^2^. This stems from limited high-level evidence and differences in approaches to stable lesions^5, 23^. Our results reinforce these concerns, demonstrating similar healing durations for successful surgical and non-operative care, with failed non-operative cases requiring substantially longer recovery.

Prior work has shown that healing can be visualized with quantitative MRI. Kajabi et al.^12^ found that T2* mapping distinguished healers from non-healers on follow-up, supporting the feasibility of ossification tracking. Our system builds on this concept, offering a clinically practical method to infer healing stage via GRE-based ossification assessment. While this study did not directly assess follow-up imaging, it establishes the foundation for structured longitudinal evaluation.

### Generalizability and Reader Reliability

Another strength of this system is its generalizability. It demonstrated substantial agreement across five independent readers, including musculoskeletal radiologists and trainees. High interrater reliability (Fleiss’ Kappa = 0.71) supports its reproducibility and clinical utility^33^. Inconsistent interpretation is a key barrier to the adoption of MRI-based OCD grading^13,16, 34^; this system resolves that issue through well-defined morphological stages and imaging correlates.

### Broader Clinical Applicability

While developed for OCD, this MRI-based staging system may have broader utility in musculoskeletal radiology. The healing of OCD follows the sequential stages of endochondral ossification – beginning with a cartilage template, initial mineralization, new bone formation and bridging, and ultimately osseous integration - mirroring the biological progression seen in fracture healing and other forms of skeletal repair^11, 35^. Gerstenfeld et al. pioneered the biological understanding of “fracture healing as a post-natal developmental process”, a perspective we now extend to OCD.

This framework challenges the enduring notion of OCD as a mechanically unstable “bony fragment”, reframing it as failure of normal endochondral ossification. The paradigm shift – from mechanical detachment to disrupted but restorable development - has implications for how OCD is diagnosed, staged and treated.

Given these shared ossification pathways, the proposed MRI-based staging system may extend beyond OCD. Short echo time GRE imaging may support assessment of fracture healing, non-union, osteotomy and osteochondral graft integration. By enhancing visualization of early bone formation, this method offers radiologists a reproducible tool for longitudinal evaluation of healing, with potential impact on clinical decision-making across orthopedic conditions.

### Limitations

This study has limitations. As a retrospective study, the staging system reflects the healing status at the time of first MRI, not the onset of disease or rate of progression. Because the initiation of ossification is unknown, early-stage lesions may be either truly early or slow-healing. Thus, treatment outcomes may depend more on the tempo of healing than on initial stage. The retrospective nature may also introduce inherent bias; however, proportional stratification across all healing stages aimed to mitigate sampling imbalance. While substantial interrater reliability was achieved, absence of serial imaging limited the ability to assess intra-subject healing trajectories over time. Although statistical power was sufficient for interrater agreement and group comparisons, the subgroup analysis—particularly for failed non-operative cases—was based on relatively small sample sizes, which may reduce precision in effect size estimates. Future prospective studies incorporating standardized follow-up imaging and larger cohorts will be essential to confirm clinical impact. Clinical outcome correlation was limited to healing endpoints rather than interim changes, and thus, the system’s potential to guide real-time treatment decisions remains to be prospectively tested.

### Conclusion

This novel MRI-based radiological staging system for OCD demonstrates substantial interrater reliability and enables assessment of healing through improved visualization of progressive ossification using short echo time GRE. By capturing sequential stages of endochondral ossification, it provides a biologically grounded framework for staging lesion maturation. Its standardized imaging approach may extend to other clinical contexts where similar ossification pathways are critical to musculoskeletal repair.

## Data Availability

All data produced in the present work are available upon reasonable request to the authors.

## Acknowledgement

This work was supported by the National Institutes of Health, including the National Institute of Arthritis and Musculoskeletal and Skin Diseases (R01 AR070020) and National Institute of Biomedical Imaging and Bioengineering (P41 EB027061). Funding sources had no role in the study or its design. We acknowledged Matthew White, Bhargavi Goduguchinta and Wendy Elvendahl for assistance in data acquisition.

## References

1. Pareek A, Sanders TL, Wu IT, Larson DR, Saris DBF, Krych AJ. Incidence of symptomatic osteochondritis dissecans lesions of the knee: a population-based study in Olmsted County. Osteoarthritis Cartilage. 2017;25(10):1663–71. Epub 2017/07/18. doi: 10.1016/j.joca.2017.07.005. PubMed PMID: 28711583; PMCID: PMC5798004.

2. Chau MM, Klimstra MA, Wise KL, Ellermann JM, Tóth F, Carlson CS, Nelson BJ, Tompkins MA. Osteochondritis Dissecans: Current Understanding of Epidemiology, Etiology, Management, and Outcomes. J Bone Joint Surg Am. 2021;103(12):1132–51. Epub 2021/06/11. doi: 10.2106/jbjs.20.01399. PubMed PMID: 34109940; PMCID: PMC8272630.

3. Laor T, Zbojniewicz AM, Eismann EA, Wall EJ. Juvenile osteochondritis dissecans: is it a growth disturbance of the secondary physis of the epiphysis? AJR Am J Roentgenol. 2012;199(5):1121–8. Epub 2012/10/26. doi: 10.2214/AJR.11.8085. PubMed PMID: 23096188.

4. Ellermann J, Johnson CP, Wang L, Macalena JA, Nelson BJ, LaPrade RF. Insights into the Epiphyseal Cartilage Origin and Subsequent Osseous Manifestation of Juvenile Osteochondritis Dissecans with a Modified Clinical MR Imaging Protocol: A Pilot Study. Radiology. 2017;282(3):798–806. Epub 2016/09/16. doi: 10.1148/radiol.2016160071. PubMed PMID: 27631413.

5. Hevesi M, Sanders TL, Pareek A, Milbrandt TA, Levy BA, Stuart MJ, Saris DBF, Krych AJ. Osteochondritis Dissecans in the Knee of Skeletally Immature Patients: Rates of Persistent Pain, Osteoarthritis, and Arthroplasty at Mean 14-Years’ Follow-Up. Cartilage. 2020;11(3):291–9. Epub 2018/07/13. doi: 10.1177/1947603518786545. PubMed PMID: 29998745; PMCID: PMC7298597.

6. Wall EJ, Milewski MD, Carey JL, Shea KG, Ganley TJ, Polousky JD, Grimm NL, Eismann EA, Jacobs JC, Jr., Murnaghan L, Nissen CW, Myer GD, Research in Osteochondritis of the Knee G, Weiss J, Edmonds EW, Anderson AF, Lyon RM, Heyworth BE, Fabricant PD, Zbojniewicz A. The Reliability of Assessing Radiographic Healing of Osteochondritis Dissecans of the Knee. Am J Sports Med. 2017;45(6):1370–5. Epub 2017/04/12. doi: 10.1177/0363546517698933. PubMed PMID: 28398084.

7. Nissen CW, Albright JC, Anderson CN, Busch MT, Carlson C, Carsen S, Chambers HG, Edmonds EW, Ellermann JM, Ellis HB, Jr., Erickson JB, Fabricant PD, Ganley TJ, Green DW, Grimm NL, Heyworth BE, Po JHH, Kocher MS, Kostyun RO, Krych AJ, Latz KH, Loveland DM, Lyon RM, Mayer SW, Meenen NM, Milewski MD, Myer GD, Nelson BJ, Nepple JJ, Nguyen JC, Pace JL, Paterno MV, Pennock AT, Perkins CA, Polousky JD, Saluan P, Shea KG, Shearier E, Tompkins MA, Wall EJ, Weiss JM, Willimon SC, Wilson PL, Wright RW, Zbojniewicz AM, Carey JL, Group R. Descriptive Epidemiology From the Research in Osteochondritis Dissecans of the Knee (ROCK) Prospective Cohort. Am J Sports Med. 2022;50(1):118–27. Epub 20211124. doi: 10.1177/03635465211057103. PubMed PMID: 34818065.

8. Toth F, Nissi MJ, Ellermann JM, Wang L, Shea KG, Polousky J, Carlson CS. Novel Application of Magnetic Resonance Imaging Demonstrates Characteristic Differences in Vasculature at Predilection Sites of Osteochondritis Dissecans. Am J Sports Med. 2015;43(10):2522–7. Epub 2015/08/20. doi: 10.1177/0363546515596410. PubMed PMID: 26286878; PMCID: PMC4766866.

9. van der Weiden GS, van Cruchten S, van Egmond N, Mastbergen SC, Husen M, Saris DBF, Custers RJH. Osteochondritis Dissecans of the Knee Associated With Mechanical Overload. Am J Sports Med. 2024;52(1):155–63. doi: 10.1177/03635465231211497. PubMed PMID: 38164681; PMCID: PMC10762890.

10. Yellin JL, Trocle A, Grant SF, Hakonarson H, Shea KG, Ganley TJ. Candidate Loci are Revealed by an Initial Genome-wide Association Study of Juvenile Osteochondritis Dissecans. Journal of pediatric orthopedics. 2017;37(1):e32–e6. Epub 2015/10/01. doi: 10.1097/BPO.0000000000000660. PubMed PMID: 26422391.

11. Morgan EF, Giacomo A, Gerstenfeld LC. Overview of Skeletal Repair (Fracture Healing and Its Assessment). Methods Mol Biol. 2021;2230:17–37. doi: 10.1007/978-1-0716-1028-2_2. PubMed PMID: 33197006.

12. Kajabi AW, Zbyn S, Johnson CP, Tompkins MA, Nelson BJ, Takahashi T, Shea KG, Marette S, Carlson CS, Ellermann JM. Longitudinal 3T MRI T(2) * mapping of Juvenile osteochondritis dissecans (JOCD) lesions differentiates operative from non-operative patients-Pilot study. J Orthop Res. 2023;41(1):150–60. Epub 20220430. doi: 10.1002/jor.25343. PubMed PMID: 35430743; PMCID: PMC9573934.

13. Andriolo L, Solaro L, Altamura SA, Carey JL, Zaffagnini S, Filardo G. Classification Systems for Knee Osteochondritis Dissecans: A Systematic Review. Cartilage. 2022;13(3):19476035221121789. doi: 10.1177/19476035221121789. PubMed PMID: 36117427; PMCID: PMC9634996.

14. Kijowski R, Blankenbaker DG, Shinki K, Fine JP, Graf BK, De Smet AA. Juvenile versus adult osteochondritis dissecans of the knee: appropriate MR imaging criteria for instability. Radiology. 2008;248(2):571–8. Epub 2008/06/13. doi: 10.1148/radiol.2482071234. PubMed PMID: 18552309.

15. Ellermann JM, Donald B, Rohr S, Takahashi T, Tompkins M, Nelson B, Crawford A, Rud C, Macalena J. Magnetic Resonance Imaging of Osteochondritis Dissecans: Validation Study for the ICRS Classification System. Academic radiology. 2016;23(6):724–9. Epub 2016/03/16. doi: 10.1016/j.acra.2016.01.015. PubMed PMID: 26976624.

16. Fabricant PD, Milewski MD, Kostyun RO, Wall EJ, Zbojniewicz AM, Research in Osteochondritis of the Knee Study G, Albright JC, Bauer KL, Carey JL, Chambers HG, Edmonds EW, Ellis HB, Ganley TJ, Green DW, Grimm NL, Heyworth BE, Kocher MS, Krych AJ, Lyon RM, Mayer SW, Nepple JJ, Nissen CW, Pennock AT, Polousky JD, Saluan P, Shea KG, Tompkins MA, Weiss J, Clifton Willimon S, Wilson PL, Wright RW, Myer GD. Osteochondritis Dissecans of the Knee: An Interrater Reliability Study of Magnetic Resonance Imaging Characteristics. Am J Sports Med. 2020;48(9):2221–9. Epub 2020/06/26. doi: 10.1177/0363546520930427. PubMed PMID: 32584594.

17. Cahill BR, Phillips MR, Navarro R. The results of conservative management of juvenile osteochondritis dissecans using joint scintigraphy. A prospective study. Am J Sports Med. 1989;17(5):601–5; discussion 5-6. Epub 1989/09/01. doi: 10.1177/036354658901700502. PubMed PMID: 2610273.

18. Robertson W, Kelly BT, Green DW. Osteochondritis dissecans of the knee in children. Curr Opin Pediatr. 2003;15(1):38–44. Epub 2003/01/25. doi: 10.1097/00008480-200302000-00007. PubMed PMID: 12544270.

19. Krause M, Hapfelmeier A, Moller M, Amling M, Bohndorf K, Meenen NM. Healing predictors of stable juvenile osteochondritis dissecans knee lesions after 6 and 12 months of nonoperative treatment. Am J Sports Med. 2013;41(10):2384–91. Epub 2013/07/24. doi: 10.1177/0363546513496049. PubMed PMID: 23876519.

20. Wall EJ, Vourazeris J, Myer GD, Emery KH, Divine JG, Nick TG, Hewett TE. The healing potential of stable juvenile osteochondritis dissecans knee lesions. J Bone Joint Surg Am. 2008;90(12):2655–64. Epub 2008/12/03. doi: 10.2106/JBJS.G.01103. PubMed PMID: 19047711; PMCID: PMC2663329.

21. Stinson ZS, Davelaar CMF, Kiebzak GM, Edmonds EW. Treatment Decisions in Pediatric Sports Medicine: Do Personal and Professional Bias Affect Decision-Making? Orthop J Sports Med. 2021;9(10):23259671211046258. Epub 20211014. doi: 10.1177/23259671211046258. PubMed PMID: 34676272; PMCID: PMC8524719.

22. Landis JR, Koch GG. An application of hierarchical kappa-type statistics in the assessment of majority agreement among multiple observers. Biometrics. 1977;33(2):363–74. PubMed PMID: 884196.

23. Edmonds EW, Polousky J. A review of knowledge in osteochondritis dissecans: 123 years of minimal evolution from Konig to the ROCK study group. Clin Orthop Relat Res. 2013;471(4):1118–26. Epub 2012/03/01. doi: 10.1007/s11999-012-2290-y. PubMed PMID: 22362466; PMCID: PMC3586043.

24. Dipaola JD, Nelson DW, Colville MR. Characterizing osteochondral lesions by magnetic resonance imaging. Arthroscopy. 1991;7(1):101–4. doi: 10.1016/0749-8063(91)90087-e. PubMed PMID: 2009106.

25. De Smet AA, Ilahi OA, Graf BK. Reassessment of the MR criteria for stability of osteochondritis dissecans in the knee and ankle. Skeletal Radiol. 1996;25(2):159–63. Epub 1996/02/01. doi: 10.1007/s002560050054. PubMed PMID: 8848747.

26. Hefti F, Beguiristain J, Krauspe R, Moller-Madsen B, Riccio V, Tschauner C, Wetzel R, Zeller R. Osteochondritis dissecans: a multicenter study of the European Pediatric Orthopedic Society. J Pediatr Orthop B. 1999;8(4):231–45. Epub 1999/10/08. PubMed PMID: 10513356.

27. Carey JL, Wall EJ, Grimm NL, Ganley TJ, Edmonds EW, Anderson AF, Polousky J, Murnaghan ML, Nissen CW, Weiss J, Lyon RM, Chambers HG, Research in OsteoChondritis of the Knee G. Novel Arthroscopic Classification of Osteochondritis Dissecans of the Knee: A Multicenter Reliability Study. Am J Sports Med. 2016;44(7):1694–8. Epub 20160406. doi: 10.1177/0363546516637175. PubMed PMID: 27159302.

28. Milewski MD, Miller PE, Gossman EC, Coene RP, Tompkins MA, Anderson CN, Bauer K, Busch MT, Carey JL, Carsen S, Chambers HG, Coene RP, Edmonds EW, Ellermann J, Ellis HB, Jr., Erickson J, Fabricant PD, Ganley TJ, Gossman EC, Green DW, Heyworth BE, Hoi Po Hui J, Kocher MS, Krych AJ, Latz K, Lyon RM, Mayer S, Milewski MD, Miller PE, Nelson BJ, Nepple JJ, Nguyen JC, Nissen CW, Lee Pace J, Paterno MV, Pennock AT, Perkins C, Polousky JD, Saluan P, Shea KG, Tompkins MA, Wall EJ, Weiss JM, Willimon C, Wilson P, Wright RW, Zbojniewicz A, Myer GD. A Simple Clinical Predictive Model for Arthroscopic Mobility of Osteochondritis Dissecans Lesions of the Knee. Am J Sports Med. 2024;52(14):3543–50. Epub 20241125. doi: 10.1177/03635465241296133. PubMed PMID: 39584729.

29. Kralik SF, Supakul N, Wu IC, Delso G, Radhakrishnan R, Ho CY, Eley KA. Black bone MRI with 3D reconstruction for the detection of skull fractures in children with suspected abusive head trauma. Neuroradiology. 2019;61(1):81–7. Epub 20181107. doi: 10.1007/s00234-018-2127-9. PubMed PMID: 30406272.

30. Low XZ, Lim MC, Nga V, Sundar G, Tan AP. Clinical application of “black bone” imaging in paediatric craniofacial disorders. Br J Radiol. 2021;94(1124):20200061. Epub 20210708. doi: 10.1259/bjr.20200061. PubMed PMID: 34233472; PMCID: PMC8764919.

31. Zbyn S, Santiago C, Johnson CP, Ludwig KD, Zhang L, Marette S, Tomplins MA, Nelson B, Metzger GJ, Carlson CS, Ellermann JM. Quantitative Evaluation of Degree of Ossification in Juvenile Osteochondritis Dissecans Lesions of the Knee with T2* Mapping. Eur J Radiol. 2021;subm. doi: 10.2106/JBJS.OA.19.00031. PubMed PMID: 32043049; PMCID: PMC6959910.

32. Ma YJ, Jerban S, Jang H, Chang D, Chang EY, Du J. Quantitative Ultrashort Echo Time (UTE) Magnetic Resonance Imaging of Bone: An Update. Front Endocrinol (Lausanne). 2020;11:567417. Epub 20200918. doi: 10.3389/fendo.2020.567417. PubMed PMID: 33071975; PMCID: PMC7531487.

33. Benomar A, Zarour E, Letourneau-Guillon L, Raymond J. Measuring Interrater Reliability. Radiology. 2023;309(3):e230492. doi: 10.1148/radiol.230492. PubMed PMID: 38085087.

34. Bexkens R, Simeone FJ, Eygendaal D, van den Bekerom MP, Oh LS, Shoulder, Elbow P. Interobserver reliability of the classification of capitellar osteochondritis dissecans using magnetic resonance imaging. Shoulder Elbow. 2020;12(4):284–93. Epub 2020/08/13. doi: 10.1177/1758573218821151. PubMed PMID: 32782483; PMCID: PMC7400717.

35. Gerstenfeld LC, Cullinane DM, Barnes GL, Graves DT, Einhorn TA. Fracture healing as a post-natal developmental process: molecular, spatial, and temporal aspects of its regulation. J Cell Biochem. 2003;88(5):873–84. doi: 10.1002/jcb.10435. PubMed PMID: 12616527.

